# Exploring risk factors associated with catastrophic health expenditure on diarrhea management in Karachi, Pakistan, 2022-2024

**DOI:** 10.1101/2025.10.16.25338206

**Authors:** Naveed Ahmed, Muhammad Tahir Yousafzai, Sonia I. Rao, Arianna Rubin Means, Patricia B. Pavlinac, Chloe Morozoff, Farah Naz Qamar

## Abstract

**Background:** Catastrophic health expenditure (CHE) is a situation where a household’s out of pocket medical payments are excessively high relative to its income, potentially leading to impoverishment. This study explores the risk factors associated with CHE in the management of diarrhea among children enrolled in Enterics for Global Health (EFGH) facilities in Karachi, Pakistan.

**Methods:** We conducted a secondary analysis of data from 1,400 children presenting with diarrhea at Pakistan EFGH facilities. We estimated households’ direct medical and non-medical costs incurred during a diarrhea episode. CHE was defined as healthcare expenditures exceeding 10% of total monthly household expenditure. Risk factors for CHE were assessed using univariate and multivariate regression models.

**Results:** CHE was observed in 9 (0.6%) households. Children experiencing severe diarrhea were 9.53 times (95% CI: 1.93-46.91, p=0.006) more likely to incur CHE compared to those with mild cases. Children with Cryptosporidium had a 7.08 times higher risk (95% CI: 1.46-34.38, p=0.015) compared to non- cryptosporidium. Children who presented at the health facility with both diarrhea and moderate or severe wasting were 5.47 and 9.71 times more likely to experience CHE compared to those without wasting (95% CI: 1.01-29.69, p=0.011 and 1.64-57.6, p=0.012) respectively. Similarly, children who were moderately underweight were 5.4 times more likely to experience CHE compared to those who were not underweight (95% CI: 1.01-29.69, p=0.043). After adjusting for diarrhea severity, the risk of CHE among households where the father’s education was limited to Quranic schooling was 8.4 times higher compared to no formal education (95% CI: 1.13-62.49, p=0.038).

**Conclusion:** Although CHE related to child diarrhea treatment is rare, it can have severe economic outcomes. Severe diarrhea, malnutrition, and *Cryptosporidium* infection were predictors of increased risk of CHE. Improving access to affordable healthcare, vaccination and nutritional programs, water, sanitation, and hygiene (WASH) practices and financial support could reduce the impact of CHE and protect vulnerable households.

## Introduction

Diarrhea is the second leading cause of death among children under five globally, claiming over half a million lives each year, which equates to 1 in 9 child deaths daily [1, 2]. Children living in low- and-middle-income countries (LMICs) are disproportionately affected, with sub-Saharan Africa and South Asia accounting for 90% of these deaths due to inadequate access to water, sanitation, and hygiene (WASH) [3–5]. In Pakistan, 74 out of every 1,000 children die annually from diarrheal diseases largely due to poor WASH practices and a fragile health system infrastructure [6]. Although diarrhea is preventable and treatable with low-cost interventions, it still results in frequent healthcare visits in LMICs [7, 8].

There are proven strategies available for the treatment and prevention of childhood diarrhea, including oral rehydration therapy, zinc supplementation, rotavirus vaccine, improvement in WASH practices, emphasis on exclusive breastfeeding and improving caregiver healthcare seeking behavior [1, 2, 9]. There has been a slight decline in cases of diarrhea with the introduction of the rotavirus vaccine, however, other pathogens continue to cause diarrhea among children [10].

Childhood diarrhea can pose a significant financial burden on families and the healthcare system, particularly in resource-limited settings. In Pakistan and other LMICs, many families incur out-of-pocket (OOP) medical payments when seeking care, which especially in lower wealth quintiles, can cause financial hardship and could be a barrier to timely access to healthcare seeking [11, 12]. The average cost of treating diarrhea varies across countries and visit characteristics, but is generally estimated to cost approximately $40 per outpatient visit and $160 per inpatient episode in LMICs, when capturing both health system and household costs (costs in 2015 USD) [9]. One study estimated the mean cost per episode of diarrhea incurred by households in Pakistan to be $6.47, with 10% of cases surpassing $10.11 (2011 USD), while another study estimated costs to range from $3.32 to $6.32 (2012 USD) depending on the pathogen [13, 14]. In Pakistan and other LMICs, state-provided healthcare services are often insufficient in providing complete coverage of medical fees, leaving families vulnerable to OOP expenditures. In addition, families often incur non-medical costs, such as transportation fees to health facilities. Given the frequency of childhood diarrhea, diarrhea treatment can become financially burdensome to both families and healthcare systems [12, 15, 16].

Catastrophic health expenditure (CHE) describes the unfortunate scenario where medical costs are so high, that they risk impoverishing a household. CHE is often estimated as OOP expenditure (direct medical and direct non-medical) for a health condition exceeding a certain threshold (generally 10% or 25%) of monthly household expenditure [8, 17, 18]. CHE disproportionately affects households with lower socioeconomic status (SES) [19–21]. Households with sufficient financial capacity can buffer medical expenses and avoid catastrophic health impacts, while in poorer households, even low levels of health spending can cause devastating financial consequences [22–24]. Data on CHE due to child diarrhea is important for policy makers’ decisions around prioritizing, upgrading and introducing new interventions to curb the disease. This study aims to estimate CHE among families of children aged under 5 years with medically attended diarrhea enrolled in the EFGH *Shigella* surveillance study in Karachi Pakistan.

## Methods

### Parent study

The Enterics for Global Health (EFGH) *Shigella* surveillance study was conducted in seven countries: Bangladesh, Kenya, Malawi, Mali, Pakistan, Peru and The Gambia. The aim of this study was to determine the incidence and consequences of medically attended diarrhea (MAD) among children aged 6-35 months. This was a facility-based MAD hybrid surveillance study conducted at selected health facilities for 24 months. The study enrolled children with diarrhea with 3 months follow-up. The full study protocol is published elsewhere [25].

### Study design

This was a secondary analysis of data obtained from the EFGH study. The aim of this analysis was to explore risk factors associated with CHE on diarrhea management in households of children enrolled in EFGH from the outpatient departments (OPDs) of six health facilities in Karachi, Pakistan. We used case report forms (CRFs) to collect information from study participants across time points. We used CRF data from enrollment, discharge and week 4 follow-up visits describing caregiver reported expenses associated with management of the diarrhea episode. Client and visit characteristics were obtained from the data treatment record CRFs (Table 1).

**Table 1:** Key cost elements and associated data sources used to measure catastrophic health expenditure in EFGH, Pakistan, 2022-2024.

| Data | Data source | Purpose |
| --- | --- | --- |
| Caregiver reported fees associated with child's episode of diarrhea | Enrollment survey, discharge survey, 4-week follow-up survey (EFGH study) | Estimating caregivers' out-of-pocket expenditures (numerator for catastrophic spending) |
| Household monthly expenditure according to wealth quintile | Household Integrated Economic Survey (HIES) 2018-19<sup>1</sup> | Estimating household monthly expenditure (denominator for catastrophic spending) |
| Threshold for catastrophic spending | Budget share approach using 10% of a household's total income or expenditure within a given period<sup>2</sup> | Estimating cut-off value for catastrophic expenditure |
| Client and visit characteristics | Clinical treatment records, enrollment survey (EFGH study) | Study covariates to determine predictors of catastrophic spending |

<sup>1</sup>Pakistan Bureau of Statistics. Household Integrated Economic Survey (HIES) 2018-19. Government of Pakistan; 2020
<sup>2</sup>Nguyen et al., 2023

### Study setting

Data of children enrolled at six EFGH study health facilities (Abbasi Shaheed Hospital, Khidmat- e-Alam Medical Centre, Sindh Government Hospital Korangi 5, Sindh Government Hospital Ibrahim Hyderi, Ali Akbar Shah, Vital Pakistan Trust (VPT) Center, and Bhains Colony VPT Center (BHC)) situated in Nazimabad and Bin-Qasim town are included in this study. These facilities are a mix of tertiary care public sector hospital and primary healthcare centers run by non- governmental organizations. All facilities serve population of low-and middle-income urban slums and provide treatment free of cost except Khidmat-e-Alam, Medical Center where minimal charges are paid by the caregiver. Description of these health facilities is published elsewhere [26].

### Study participants

Children aged 6-35 months who visited OPDs of the EFGH health facilities were eligible for enrollment. Children who presented with the complaint of diarrhea, defined as more than three stools in the last 24 hours, living in the defined catchment area with plans to remain in the catchment area for the next four months were approached for possible enrollment in the study. Details of the exclusion criteria have been reported elsewhere. [27].

### Study outcome

The outcome of this was the occurrence of catastrophic health expenditure which was defined using the budget share approach, also called the basic approach used by the World Bank to ascertain CHE [28]. The budget share approach defines CHE as OOP expenditures related to a health condition exceeding a certain proportion of total household income or expenditure within a given period [18]. Specifically, we defined CHE as meeting or exceeding a threshold of 10% of monthly expenditures, which had most commonly been used in the context of low- and middle-income settings [29].

Diarrhea was defined as the passage of 3 or more loose or liquid stools per day (or more frequent passage than normal for the individual), and MAD was defined as any diarrheal episode for which medical care is sought at an EFGH enrollment site. We also assessed direct medical cost, which encompassed expenses pertaining to diagnosis and treatment of a child with MAD, either in outpatient or inpatient settings. In our analysis, we included the portion of direct medical costs paid by caregivers which include clinical registration fees, lab tests or diagnostics, medications and other medical supplies purchased in or outside of the facility. We also included direct non-medical costs which included the cost of transportation to reach the healthcare facility, food or accommodation while children were being treated, or other non-medical fees spent related to the visit (e.g., childcare for other children).

### Data collection

We primarily utilized data collected through the EFGH study related to child demographic and anthropometry indicators, as well as visit costs. During a child’s enrollment visit at the health facility, a study physician conducted a physical examination, collecting anthropometry measurements including child length, weight and mid-upper arm circumference. A caregiver interview was first conducted during the child’s enrollment visit, to collect information on socio- demographic characteristics, health seeking behaviors, and cost to access healthcare facility and treatment. Caregiver interviews also included a simplified asset questionnaire, adapted from the Demographic Health Survey wealth index form, to estimate the household’s wealth quintile [30]. A stool sample was collected at baseline to test for pathogens. Additional data on direct medical and direct non-medical costs were collected at 4-week follow-up visits, to capture new costs incurred after leaving the health facility.

The EFGH study did not collect information on household incomes or expenditures, therefore, we estimated household expenditures from the government of Pakistan’s Household Integrated Economic Survey (HIES) 2018-2019 data, Pakistan Bureau of Statistics [31, 32]. Specifically, we extracted monthly household expenditure per wealth quintile from Sindh province.

### Statistical Analysis

We determined CHE status per child enrolled in the EFGH study. To do so, we first summed caregiver-reported costs to calculate total OOP fees per child visit. Each child visit was also assigned a monthly household expenditure from Pakistan’s HIES, matched based on their wealth quintile (as measured by a wealth index). All costs were collected in Pakistani Rupees, and for comparability, were inflated to 2023 currency using Gross Domestic Product price deflators from the World Bank. Per child-visit, we calculated the proportion of monthly household expenditure spent on OOP expenditure on the management of diarrhea. Cases where OOP was 10% or greater were determined to be CHE.

We considered the following risk factors for CHE: type of health facility, age and sex of the child, caregiver’s age, education status of child’s parents, number of children in house, healthcare seeking prior to enrollment in EFGH, travel time to clinic, type of pathogen and diarrhea severity. Descriptive statistics on risk factors are presented using means and standard deviations for continuous variables and counts and proportions for categorical variables. Risk ratios (RR) and 95% confidence intervals (CI) were used to describe the association between risk factors and catastrophic expenditure on diarrhea.

To determine potential predictors of CHE we constructed a regression model with generalized estimating equation (GEE) approach and poison distribution with log link, and clustering per child (to account for children with multiple diarrheal episodes). The binary dependent variable was whether the diarrheal episode resulted in CHE. We first estimated unadjusted models for each predictor, followed by multivariate models adjusting for diarrhea severity.

### Sensitivity analyses

We ran sensitivity analyses to test the effect of key analytical decisions on risk of CHE. To test the effect of the CHE definition on our results, we tested monthly household expenditure spent on the diarrheal episode at 8% and 5% threshold, as opposed to the standard 10% threshold.

### Ethical Considerations

The parent study is registered with ClinicalTrials.gov (NCT06047821). All materials including consent forms and protocol are approved from Ethics Review Committee (ERC), Aga Khan University. All children enrolled in the study received informed consent from caregivers.

## Results

### Child health, caregiver characteristics and pathogen distribution

A total of 1,400 children were included in this analysis. Of the total, 646 (46.1%) of children were 12-23 months old and 53.2% were male. The *Shigella* isolation rate among enrolled children was 25.3% followed by *Enterotoxigenic Escherichia coli* (ETEC) at 12.0%, rotavirus 8.5% and *Cryptosporidium* at 4.4%. Approximately one third of children were stunted, wasted or underweight at presentation at the health facility. The mean age of the caregivers was 27.12 ±

5.61 years. Households with male children, children with severe diarrhea and those who presented to primary healthcare facilities were more common in the CHE category (Table 2). CHE was rare, observed in 9 (0.6%) households. The largest medical and non-medical costs incurred on the management of an episode of diarrhea were drugs with mean cost of 55.9 Pakistani rupee (PKR) followed by the transportation 53.6 PKR respectively.

**Table 2:**
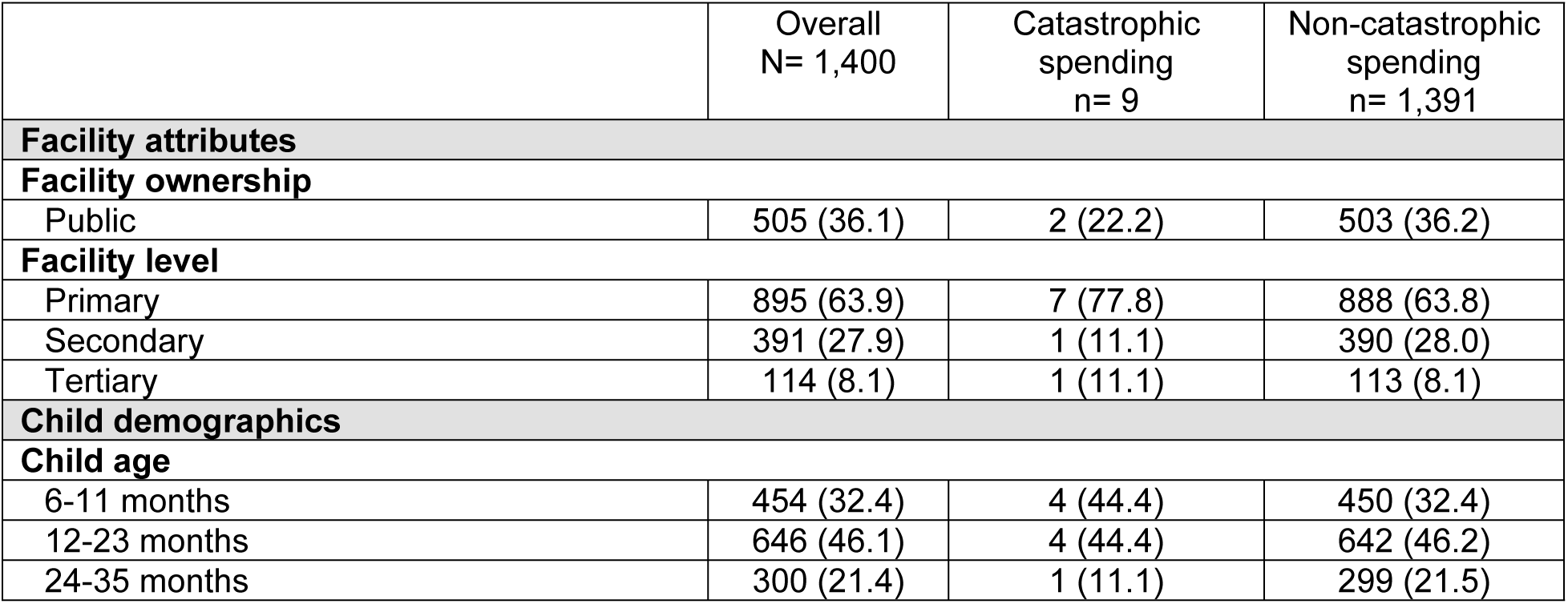
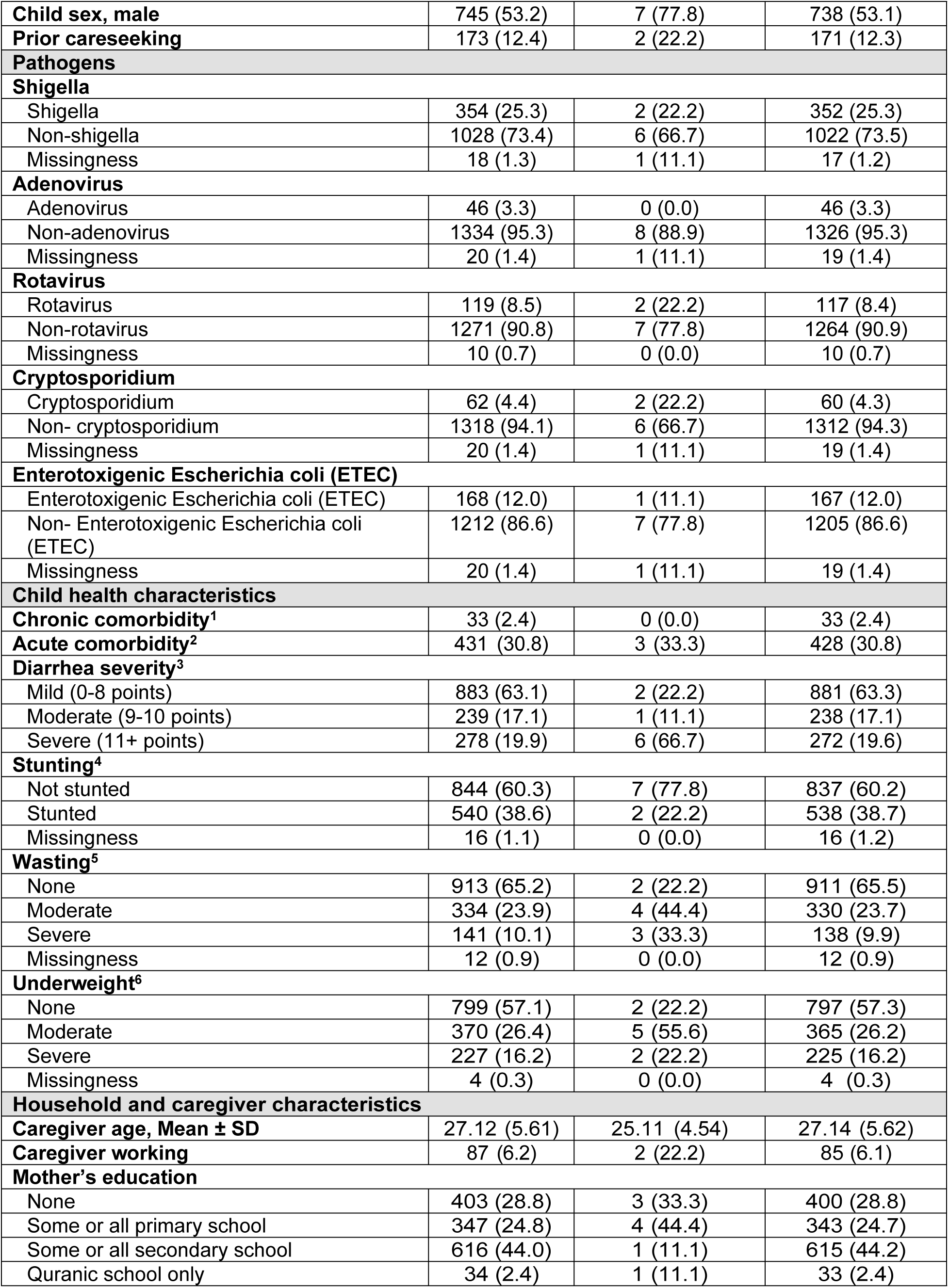
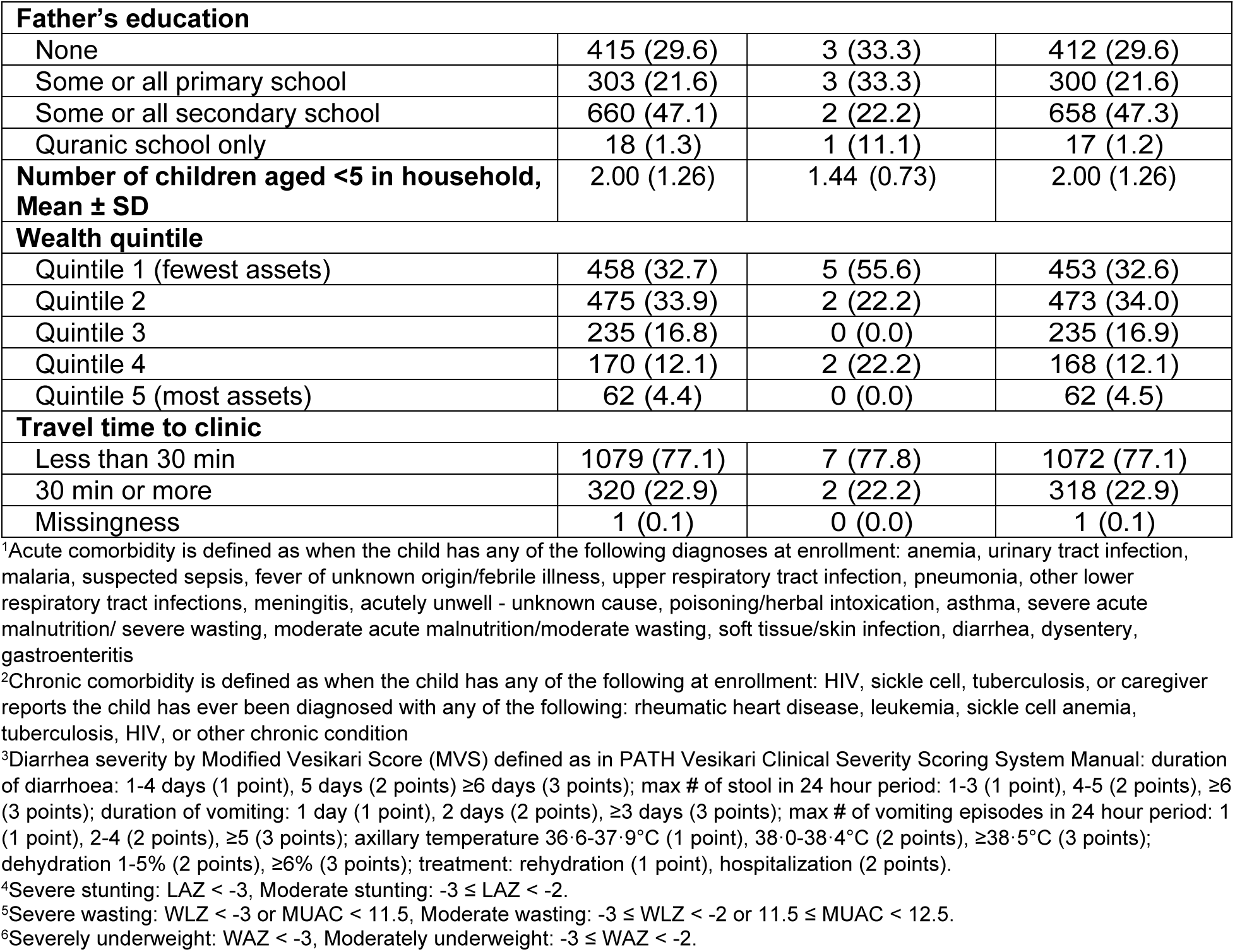
Demographic characteristics of children aged 6-35 months among households of children with catastrophic health expenditure on diarrhea care seeking in EFGH Pakistan Site, 2022-2024.

|  | Overall<br>N= 1,400 | Catastrophic<br>spending<br>n= 9 | Non-catastrophic<br>spending<br>n= 1,391 |
| --- | --- | --- | --- |
| **Facility attributes** |  |  |  |
| **Facility ownership** |  |  |  |
| Public | 505 (36.1) | 2 (22.2) | 503 (36.2) |
| **Facility level** |  |  |  |
| Primary | 895 (63.9) | 7 (77.8) | 888 (63.8) |
| Secondary | 391 (27.9) | 1 (11.1) | 390 (28.0) |
| Tertiary | 114 (8.1) | 1 (11.1) | 113 (8.1) |
| **Child demographics** |  |  |  |
| **Child age** |  |  |  |
| 6-11 months | 454 (32.4) | 4 (44.4) | 450 (32.4) |
| 12-23 months | 646 (46.1) | 4 (44.4) | 642 (46.2) |
| 24-35 months | 300 (21.4) | 1 (11.1) | 299 (21.5) |
| **Child sex, male** | 745 (53.2) | 7 (77.8) | 738 (53.1) |
| **Prior careseeking** | 173 (12.4) | 2 (22.2) | 171 (12.3) |
| **Pathogens** |  |  |  |
| **Shigella** |  |  |  |
| Shigella | 354 (25.3) | 2 (22.2) | 352 (25.3) |
| Non-shigella | 1028 (73.4) | 6 (66.7) | 1022 (73.5) |
| Missingness | 18 (1.3) | 1 (11.1) | 17 (1.2) |
| **Adenovirus** |  |  |  |
| Adenovirus | 46 (3.3) | 0 (0.0) | 46 (3.3) |
| Non-adenovirus | 1334 (95.3) | 8 (88.9) | 1326 (95.3) |
| Missingness | 20 (1.4) | 1 (11.1) | 19 (1.4) |
| **Rotavirus** |  |  |  |
| Rotavirus | 119 (8.5) | 2 (22.2) | 117 (8.4) |
| Non-rotavirus | 1271 (90.8) | 7 (77.8) | 1264 (90.9) |
| Missingness | 10 (0.7) | 0 (0.0) | 10 (0.7) |
| **Cryptosporidium** |  |  |  |
| Cryptosporidium | 62 (4.4) | 2 (22.2) | 60 (4.3) |
| Non- cryptosporidium | 1318 (94.1) | 6 (66.7) | 1312 (94.3) |
| Missingness | 20 (1.4) | 1 (11.1) | 19 (1.4) |
| **Enterotoxigenic Escherichia coli (ETEC)** |  |  |  |
| Enterotoxigenic Escherichia coli (ETEC) | 168 (12.0) | 1 (11.1) | 167 (12.0) |
| Non- Enterotoxigenic Escherichia coli (ETEC) | 1212 (86.6) | 7 (77.8) | 1205 (86.6) |
| Missingness | 20 (1.4) | 1 (11.1) | 19 (1.4) |
| **Child health characteristics** |  |  |  |
| **Chronic comorbidity<sup>1</sup>** | 33 (2.4) | 0 (0.0) | 33 (2.4) |
| **Acute comorbidity<sup>2</sup>** | 431 (30.8) | 3 (33.3) | 428 (30.8) |
| **Diarrhea severity<sup>3</sup>** |  |  |  |
| Mild (0-8 points) | 883 (63.1) | 2 (22.2) | 881 (63.3) |
| Moderate (9-10 points) | 239 (17.1) | 1 (11.1) | 238 (17.1) |
| Severe (11+ points) | 278 (19.9) | 6 (66.7) | 272 (19.6) |
| **Stunting<sup>4</sup>** |  |  |  |
| Not stunted | 844 (60.3) | 7 (77.8) | 837 (60.2) |
| Stunted | 540 (38.6) | 2 (22.2) | 538 (38.7) |
| Missingness | 16 (1.1) | 0 (0.0) | 16 (1.2) |
| **Wasting<sup>5</sup>** |  |  |  |
| None | 913 (65.2) | 2 (22.2) | 911 (65.5) |
| Moderate | 334 (23.9) | 4 (44.4) | 330 (23.7) |
| Severe | 141 (10.1) | 3 (33.3) | 138 (9.9) |
| Missingness | 12 (0.9) | 0 (0.0) | 12 (0.9) |
| **Underweight<sup>6</sup>** |  |  |  |
| None | 799 (57.1) | 2 (22.2) | 797 (57.3) |
| Moderate | 370 (26.4) | 5 (55.6) | 365 (26.2) |
| Severe | 227 (16.2) | 2 (22.2) | 225 (16.2) |
| Missingness | 4 (0.3) | 0 (0.0) | 4 (0.3) |
| **Household and caregiver characteristics** |  |  |  |
| **Caregiver age, Mean ± SD** | 27.12 (5.61) | 25.11 (4.54) | 27.14 (5.62) |
| **Caregiver working** | 87 (6.2) | 2 (22.2) | 85 (6.1) |
| **Mother's education** |  |  |  |
| None | 403 (28.8) | 3 (33.3) | 400 (28.8) |
| Some or all primary school | 347 (24.8) | 4 (44.4) | 343 (24.7) |
| Some or all secondary school | 616 (44.0) | 1 (11.1) | 615 (44.2) |
| Quranic school only | 34 (2.4) | 1 (11.1) | 33 (2.4) |
| **Father's education** |  |  |  |
| None | 415 (29.6) | 3 (33.3) | 412 (29.6) |
| Some or all primary school | 303 (21.6) | 3 (33.3) | 300 (21.6) |
| Some or all secondary school | 660 (47.1) | 2 (22.2) | 658 (47.3) |
| Quranic school only | 18 (1.3) | 1 (11.1) | 17 (1.2) |
| **Number of children aged <5 in household, Mean ± SD** | 2.00 (1.26) | 1.44 (0.73) | 2.00 (1.26) |
| **Wealth quintile** |  |  |  |
| Quintile 1 (fewest assets) | 458 (32.7) | 5 (55.6) | 453 (32.6) |
| Quintile 2 | 475 (33.9) | 2 (22.2) | 473 (34.0) |
| Quintile 3 | 235 (16.8) | 0 (0.0) | 235 (16.9) |
| Quintile 4 | 170 (12.1) | 2 (22.2) | 168 (12.1) |
| Quintile 5 (most assets) | 62 (4.4) | 0 (0.0) | 62 (4.5) |
| **Travel time to clinic** |  |  |  |
| Less than 30 min | 1079 (77.1) | 7 (77.8) | 1072 (77.1) |
| 30 min or more | 320 (22.9) | 2 (22.2) | 318 (22.9) |
| Missingness | 1 (0.1) | 0 (0.0) | 1 (0.1) |

<sup>1</sup>Acute comorbidity is defined as when the child has any of the following diagnoses at enrollment: anemia, urinary tract infection, malaria, suspected sepsis, fever of unknown origin/febrile illness, upper respiratory tract infection, pneumonia, other lower respiratory tract infections, meningitis, acutely unwell - unknown cause, poisoning/herbal intoxication, asthma, severe acute malnutrition/ severe wasting, moderate acute malnutrition/moderate wasting, soft tissue/skin infection, diarrhea, dysentery, gastroenteritis
<sup>2</sup>Chronic comorbidity is defined as when the child has any of the following at enrollment: HIV, sickle cell, tuberculosis, or caregiver reports the child has ever been diagnosed with any of the following: rheumatic heart disease, leukemia, sickle cell anemia, tuberculosis, HIV, or other chronic condition
<sup>3</sup>Diarrhea severity by Modified Vesikari Score (MVS) defined as in PATH Vesikari Clinical Severity Scoring System Manual: duration of diarrhoea: 1-4 days (1 point), 5 days (2 points) ≥ 6 days (3 points); max # of stool in 24 hour period: 1-3 (1 point), 4-5 (2 points), ≥ 6 (3 points); duration of vomiting: 1 day (1 point), 2 days (2 points), ≥ 3 days (3 points); max # of vomiting episodes in 24 hour period: 1 (1 point), 2-4 (2 points), ≥ 5 (3 points); axillary temperature 36.6-37.9°C (1 point), 38.0-38.4°C (2 points), ≥ 38.5°C (3 points); dehydration 1-5% (2 points), ≥ 6% (3 points); treatment: rehydration (1 point), hospitalization (2 points).
<sup>4</sup>Severe stunting: LAZ < -3, Moderate stunting: -3 ≤ LAZ < -2.
<sup>5</sup>Severe wasting: WLZ < -3 or MUAC < 11.5, Moderate wasting: -3 ≤ WLZ < -2 or 11.5 ≤ MUAC < 12.5.
<sup>6</sup>Severely underweight: WAZ < -3, Moderately underweight: -3 ≤ WAZ < -2.

### Factors associated with CHE

In the univariate model we tested 19 risk factors for CHE and found that severe diarrhea, *Cryptosporidium* pathogen, moderate and severe wasting, and moderate underweight were significantly associated with CHE (Table 3). After adjusting for diarrhea severity in the multivariate model, we found moderate wasting was no longer significantly associated with CHE. Father’s education limited to Quranic school only was significantly associated with CHE in multivariate analysis but not in the univariate model.

**Table 3:** Unadjusted and adjusted analysis of factors associated with catastrophic health expenditure on diarrhea care seeking among households of children aged 6-35 months in EFGH Pakistan Site, 2022-2024.

|  | Unadjusted RR<br>estimate (95% CI) | p-value | <sup>1</sup>Adjusted RR<br>estimate (95% CI) | p-value |
| --- | --- | --- | --- | --- |
| **Facility attributes** |  |  |  |  |
| **Facility ownership** |  |  |  |  |
| Private | Ref | Ref | Ref | Ref |
| Public | 0.51 (0.11-2.43) | 0.395 | 0.36 (0.08-1.64) | 0.185 |
| **Facility level** |  |  |  |  |
| Primary | Ref | Ref | Ref | Ref |
| Secondary | 0.33 (0.04-2.65) | 0.295 | 0.23 (0.03-1.8) | 0.161 |
| Tertiary | 1.12 (0.14-9.04) | 0.914 | 0.79 (0.1-6.1) | 0.825 |
| **Child demographics** |  |  |  |  |
| **Child sex** |  |  |  |  |
| Female | Ref | Ref | Ref | Ref |
| Male | 3.08 (0.64-14.76) | 0.160 | 3.11 (0.65-14.84) | 0.155 |
| **Child age** |  |  |  |  |
| 6-11 months | Ref | Ref | Ref | Ref |
| 12-23 months | 0.7 (0.18-2.79) | 0.617 | 0.82 (0.22-3.04) | 0.766 |
| 24-35 months | 0.38 (0.04-3.37) | 0.384 | 0.6 (0.06-5.61) | 0.652 |
| **Prior careseeking** |  |  |  |  |
| No | Ref | Ref | Ref | Ref |
| Yes | 2.03 (0.42-9.67) | 0.375 | 1.58 (0.31-8.01) | 0.581 |
| **Pathogen** |  |  |  |  |
| **Shigella** |  |  |  |  |
| Non-Shigella | Ref | Ref | Ref | Ref |
| Shigella | 0.97 (0.2-4.77) | 0.968 | 0.99 (0.2-4.86) | 0.989 |
| **Rotavirus** |  |  |  |  |
| Non-rotavirus | Ref | Ref | Ref | Ref |
| Rotavirus | 3.05 (0.64-14.51) | 0.161 | 2.15 (0.48-9.67) | 0.319 |
| **Cryptosporidium** |  |  |  |  |
| Non- cryptosporidium | Ref | Ref | Ref | Ref |
| Cryptosporidium | 7.08 (1.46-34.38) | **0.015** | 6.51 (1.31-32.43) | **0.022** |
| **Enterotoxigenic Escherichia coli (ETEC)** |  |  |  |  |
| Non- Enterotoxigenic Escherichia coli (ETEC) | Ref | Ref | Ref | Ref |
| Enterotoxigenic Escherichia coli (ETEC) | 1.03 (0.13-8.31) | 0.977 | 0.78 (0.1-6.09) | 0.810 |
| **Child health characteristics** |  |  |  |  |
| **Chronic comorbidity** |  |  |  |  |
| No | Ref | Ref | Ref | Ref |
| Yes | 0 (0-0) | 0.000 | 0 (0-0) | 0.000 |
| **Acute comorbidity** |  |  |  |  |
| No | Ref | Ref | Ref | Ref |
| Yes | 1.12 (0.28-4.47) | 0.868 | 0.71 (0.16-3.05) | 0.643 |
| **Diarrhea severity** |  |  |  |  |
| Mild | Ref | Ref | Ref | Ref |
| Moderate | 1.85 (0.17-20.26) | 0.616 | NA | NA |
| Severe | 9.53 (1.93-46.91) | **0.006** | -- | -- |
| **Wasting** |  |  |  |  |
| None | Ref | Ref | Ref | Ref |
| Moderate | 5.47 (1.01-29.69) | **0.049** | 4.5 (0.79-25.63) | 0.090 |
| Severe | 9.71 (1.64-57.6) | **0.012** | 8.21 (1.3-51.79) | **0.025** |
| **Stunting** |  |  |  |  |
| Not Stunned | Ref | Ref | Ref | Ref |
| Stunted | 0.45 (0.09-2.14) | 0.314 | 0.55 (0.12-2.47) | 0.439 |
| **Underweight** |  |  |  |  |
| None | Ref | Ref | Ref | Ref |
| Moderate | 5.4 (1.05-27.68) | **0.043** | 5.66 (1.12-28.68) | **0.036** |
| Severe | 3.52 (0.5-24.85) | 0.207 | 3.58 (0.51-25.25) | 0.201 |
| **Household and caregiver characteristics** |  |  |  |  |
| **Caregiver age (years)** | 0.93 (0.82-1.05) | 0.233 | 0.93 (0.82-1.05) | 0.235 |
| **Caregiver work** |  |  |  |  |
| Not Working | Ref | Ref | Ref | Ref |
| Working | 4.31 (0.91-20.47) | 0.066 | 3.67 (0.82-16.4) | 0.089 |
| **Mother education** |  |  |  |  |
| None | Ref | Ref | Ref | Ref |
| Some or all primary school | 1.55 (0.35-6.87) | 0.565 | 1.35 (0.3-5.99) | 0.696 |
| Some or all secondary school | 0.22 (0.02-2.09) | 0.187 | 0.2 (0.02-1.92) | 0.164 |
| Quranic school only | 3.95 (0.42-36.94) | 0.228 | 4.68 (0.55-39.67) | 0.157 |
| **Father education** |  |  |  |  |
| None | Ref | Ref | Ref | Ref |
| Some or all primary school | 1.37 (0.28-6.74) | 0.699 | 1.23 (0.25-6.17) | 0.801 |
| Some or all secondary school | 0.42 (0.07-2.5) | 0.340 | 0.39 (0.07-2.36) | 0.306 |
| Quranic school only | 7.68 (0.84-70.28) | 0.071 | 8.4 (1.13-62.49) | **0.038** |
| **Number of children <5 in household** | 0.5 (0.2-1.25) | 0.138 | 0.53 (0.21-1.32) | 0.170 |
| **Travel time to clinic** |  |  |  |  |
| <30 min | Ref | Ref | Ref | Ref |
| 30 min or more | 0.96 (0.2-4.61) | 0.963 | 0.82 (0.18-3.77) | 0.799 |
| **Wealth quintile** |  |  |  |  |
| Quintile 1 (fewest assets) | Ref | Ref | Ref | Ref |
| Quintile 2 | 0.39 (0.08-1.98) | 0.253 | 0.42 (0.08-2.16) | 0.298 |
| Quintile 3 | 0 (0-0) | 0.000 | 0 (0-0) | 0.000 |
| Quintile 4 | 1.08 (0.21-5.5) | 0.928 | 1.16 (0.22-6) | 0.862 |
| Quintile 5 (most assets) | 0 (0-0) | 0.000 | 0 (0-0) | 0.000 |

CI: 95% confidence interval, Significant RR is presented in bold
<sup>1</sup>All variables were adjusted for diarrhea severity

Compared to mild diarrhea cases, households with children experiencing severe diarrhea were estimated to be 9.53 times more likely to experience CHE (95% CI: 1.93- 46.91, p=0.006) and children with moderate diarrhea 1.85 times more likely (95% CI: 0.17-20.26, p=0.616). The risk of CHE among children with *Cryptosporidium* (as compared to non- *Cryptosporidium*) was estimated to be 7.08 times (95% CI: 1.46-34.38, p=0.015); and similarly, 6.51 times (95% CI: 1.31-32.43, p=0.022) more likely after adjusting for diarrhea severity.

Compared to children with no wasting, children with moderate wasting who experienced diarrhea were 5.47 times more likely to experience CHE (95% CI: 1.01-29.69, p=0.049) and those with severe wasting were 9.71 times more likely to experience CHE (95% CI: 1.64-57.6, p=0.012). After adjusting for diarrhea severity, the risk of CHE among children with severe wasting decreased slightly to 8.21 (95% CI: 1.3-51.79, p= 0.025) though remained significant (Table 3). Moderately underweight children who experienced diarrhea were 5.4 times more likely to experience CHE (95% CI: 1.05-27.68, p=0.043). After adjusting for diarrhea severity, the risk slightly increased to 5.66 and also remained significant (95% CI: 1.12-28.68, p= 0.036).

Children whose father’s education was limited to Quranic school only were 7.68 times more likely to experience CHE (95% CI: 0.84-70.28) compared to father with no formal education. After adjusting for diarrhea severity, the risk of CHE increased significantly to 8.4 times likely (95% CI: 1.13-62.49, p=0.038).

### Sensitivity analysis of CHE thresholds

When testing the effect of the CHE threshold in sensitivity analyses, we found that 12 households experienced CHE when using a threshold of 8% of monthly household expenditures spent on the diarrheal episode, while 17 households experienced CHE when the threshold was set to 5%. Having severe diarrhea remained significantly associated with CHE when using a 5% and 8% threshold. Households with children positive for *Cryptosporidium* pathogen and severe wasting were associated with CHE at the 8% threshold, but not at the 5% threshold. Chronic comorbidity and stunting were significantly associated with CHE at 5% threshold. See S1 appendix table for further details on sensitivity analysis.

## Discussion

This study explored risk factors associated with CHE on the management of diarrhea in EFGH facilities in Karachi, Pakistan. Besides affecting household financial stability, CHE can lead to poorer health outcomes, especially for the poorest, who face both serious health risks and financial hardship [12]. Our study highlighted that CHE is rare and occurred in 0.6% of the households at a threshold of 10%. The risk of CHE increased among households of children with severe diarrhea, cryptosporidium infection and compromised nutritional status. Antibiotics and transportation were the main drivers of CHE. The EFGH study sites in Pakistan are a mix of public, private and not-for-profit, therefore, the costs of the treatment may be different between sites. All the patients sought outpatient care, and none sought inpatient care, and a minimal number of patients enrolled with severe disease.

In our study, severe diarrhea and compromised nutritional status were significantly associated with an increased risk of CHE. The vulnerability and associated high cost may be due to the interplay of these factors, as poor nutritional status and underlying health conditions increase susceptibility to diarrheal disease, resulting in more frequent healthcare visits and higher medical costs leading to CHE. This aligns with findings from Mozambique, where children with diarrhea and additional health impairments, including wasting, experienced prolonged hospital stays leading to higher medical costs [33]. Our findings also align with studies by Sultana R et al. (2021) and Memirie et al. (2017), that reported higher expenditure on severe diarrhea among children leading to financial catastrophe and instability among household of low income communities [34, 35]. While wasting is partially linked to diarrhea severity and age, it also appears to independently increase the risk of CHE [33]. *Cryptosporidium* infection was significantly associated with CHE in our study. A study conducted in three developing countries reported higher treatment cost for *cryptosporidium* infection and a consequent significant cost impact on a household [36].

In our study even though the risk of CHE was rare, it still occurred in 0.6% of the households that fall in the third quintile of monthly expenditure. A modeling study reported the risk of CHE among households to be 0.1% overall and 0.4% among households at 10% threshold in income quintile one in 34 LMICs [12]. The discordance of CHE risk could be because the services in this modeling study were provided at the primary healthcare level, however, in our study a significant number of children sought care at tertiary care hospitals indicating a more severe disease. In contrast to our study, a study from Bangladesh involving 899 households reported that 47% incurred CHE at the 10% threshold of household consumption expenditure when seeking care for childhood diarrhea, with 14% spending more than 25% of their monthly resources on treatment [8]. The reason for higher occurrence of CHE in this study could be because almost 65% of the patients were hospitalized for inpatient care. In comparison, in our study, none of the patients were hospitalized for care. While CHE is rare according to traditional definitions, it’s possible that households face greater financial risk due to loss of income while caring for their sick child. In the main EFGH study [37], the indirect cost accounted for 18% of the total expenditure on the management of *Shigella* MAD in EFGH Pakistan sites, indicating a substantial burden on the families and a risk to CHE.

Our study showed low occurrence of CHE in EFGH sites in Pakistan, however, it could be more common in other areas. This may be because the treatment is provided free of cost in all EFGH study sites in Pakistan, except one. Furthermore, a majority of participants resided in close vicinity with minimal transportation cost of accessing these facilities. In the main EFGH study [37], Pakistan had the lowest household costs compared to other countries; by contrast, in other EFGH sites, the costs were higher and transportation was the primary driver of costs when seeking care for *Shigella* MAD, substantially contributing to financial burden and potentially leading to CHE. Therefore, our study population may not represent the entire population of Pakistan. In Pakistan the spending on healthcare is mostly through OOP to cover direct medical and non-medical costs [38]. Moreover, the proportion of households experiencing CHE related to any illness in the most deprived quintile (Q1) is reported to have progressed from 8.3% (2007-08) to 13.7% (2018-19) at 10% threshold [38, 39]. Therefore, preventing the occurrence of CHE is crucial because it can push families into poverty, force them to cut back on essential needs like food and education, and create long-term financial instability [38].

There are a few important limitations to this study. The EFGH study did not capture actual income or expenditure of the household, therefore, a proxy country level data [31] source on household expenditure by wealth quintile was used to document the results. The study used a 10% threshold as the primary definition of CHE, but results from sensitivity analyses applying 8% and 5% thresholds highlighted additional financial risks, particularly among children with chronic health conditions where repeated clinic visits can increase the risk of ongoing CHE for families. These results indicate that the risk of financial catastrophe is sensitive to how CHE is defined, underscoring the need for policymakers to consider multiple thresholds when designing social protection strategies.

## CONCLUSION

Our study highlights that while CHE is rare within EFGH facilities in Pakistan, it remains a significant risk for vulnerable populations, particularly those with severe diarrhea, poor nutritional status, and low paternal education. Among direct medical and direct non-medical costs, antibiotics and transportation were the main drivers of CHE. Strengthening financial risk protection, promoting preventive healthcare, and ensuring equitable access to treatment can help reduce the impact of CHE. Integrating preventive measures such as improved sanitation, nutrition support, and vaccination programs can further mitigate the economic impact of diarrheal diseases in Pakistan and other LMICs.

## Data Availability

The EFGH statistical analysis plan (https://clinicaltrials.gov/study/NCT06047821) and study protocol (https://academic.oup.com/ofid/issue/11/Supplement_1) were made publicly available. The datasets were deidentified and anonymized and will be publicly available upon publication of the manuscript.

https://academic.oup.com/ofid/issue/11/Supplement_1

## Acknowledgment

The authors thank the children who participated in this study and their families, and the dedicated physicians, nurses, scientists and staff at each study site for their dedication and outstanding performance of clinical and laboratory study activities. The authors also thank Ms. Beth A. Tippett Barr from Nyanja Health Research Institute who facilitated and contributed to the EFGH Manuscript Writing Cohort Program.

## Supporting information

**S1 Table.** Sensitivity analysis of factors associated with CHE

